# SARS-CoV-2 cross-reactive B-cells outnumber seasonal coronavirus spike-specific clones at the end of the COVID-19 pandemic

**DOI:** 10.1101/2025.09.28.25336743

**Authors:** Cristina Gonzalez Lopez, Muriel Aguilar-Bretones, Jingshu Zhang, Batuhan Bekki, Petra van den Doel, Eric C. van Gorp, P. Hugo M. van der Kuy, Bart L. Haagmans, Corine H. GeurtsVanKessel, Marion P.G. Koopmans, Rory D. de Vries, Marit J. van Gils, Gijsbert P. van Nierop

**Affiliations:** Department of Viroscience, Erasmus University Medical Center, Rotterdam, The Netherlands; Department of Hospital Pharmacy, Erasmus University Medical Center, Rotterdam, the Netherlands; Department of Medical Microbiology and Infection Prevention, Amsterdam University Medical Center, The Netherlands

**Keywords:** Seasonal coronaviruses, COVID-19, Monoclonal antibodies, B-cells, S2 domain

## Abstract

B-cell responses towards seasonal human coronaviruses (sHCoVs), particularly OC43, impacted those towards SARS-CoV-2 due to immune imprinting in severe COVID-19 patients. However, little is known on how widespread SARS-CoV-2 circulation and COVID-19 vaccination campaigns over the course of the pandemic affected immunity towards sHCoVs in the general population.

To explore potential differences in immune recognition of sHCoVs in immunocompetent adults, we compared two cross-sectional cohorts: one sampled between 2018 and 2019 (pre-pandemic), the other at the end of the pandemic (February - March 2023). We compared serum IgG and IgA titers, antibody cross-reactivity patterns at the clonal level, and specificity towards Spike (S) domains for all sHCoVs and dominant SARS-CoV-2 variants by B-cell analysis. Subsequently, we determined the OC43 neutralization potential of sera and monoclonal antibodies targeting different S domains.

In pre-pandemic individuals, SARS-CoV-2-reactive antibody and B-cell levels, and sHCoV/SARS-CoV-2 cross-reactivity were negligible. IgA and IgG reactivity against the S of sHCoVs was distributed over spike domain 1 and 2 (S1, S2). In end-pandemic donors, SARS-CoV-2-specific immune responses strongly dominated and the majority of sHCoV reactive clones cross-reacted with SARS-CoV-2. The SARS-CoV-2/sHCoVs cross-reactive clones accounted for higher NL63 S1- and OC43 S2-specific B-cell frequencies and matched higher serum antibody titers. For OC43, the immunodominance of SARS-CoV-2/OC43 cross-reactive IgG B-cells resulted in a strong bias towards S2. Serum OC43 neutralization titers were higher in end-pandemic donors and correlated with OC43 S1 and S2-specific IgG titers and reactive B-cells frequencies. However, the SARS-CoV-2/OC43 S2 cross-reactive IgG clones did not independently correlate with OC43 neutralization titers.

We conclude that the establishment of SARS-CoV-2-specific immune responses altered responses to sHCoVs in our cohort, particularly for OC43 and NL63. This could have implications for the immune protection and offers insights for the development of pan-coronavirus treatments and vaccines.

## INTRODUCTION

Population immunity towards severe acute respiratory syndrome coronavirus 2 (SARS-CoV-2) was induced by a mix of widespread circulation of the virus and coronavirus disease 2019 (COVID-19) vaccination campaigns. Low levels of pre-existing SARS-CoV-2 reactive serum antibodies and B-cells have been detected in naïve individuals, likely induced by prior infections with seasonal human coronaviruses (sHCoVs)^1,2^.

Coronaviruses^2–4^ are classified in alpha-coronaviruses (alpha-CoV), which include sHCoVs 229E and NL63, and beta-coronaviruses (beta-CoV), including sHCoVs OC43 and HKU1, all of which generally cause common cold symptoms and are repeatedly encountered from childhood onwards^2–4^. However, the beta-CoV also include the more recently emerged and more pathogenic Middle East respiratory syndrome-related Coronavirus (MERS-CoV), SARS-CoV, and SARS-CoV-2. Alpha- and beta-CoVs share a variable degree of sequence and structural homology with SARS-CoV-2, which is highest with the beta-CoV^5^. Coronaviruses have a Spike glycoprotein (S) that governs attachment to and fusion with the target cell. S protrudes from the viral particle in trimeric form and consists of the membrane-proximal domain S2, containing the fusion apparatus, and the outer domain S1, containing the receptor binding domain (RBD). Homology between the S proteins of sHCoVs, SARS-CoV-2 and its variants is higher in the S2 domain than in S1, where most variability is observed in the RBD^6^.

The structural homology between SARS-CoV-2 and other coronaviruses translates into antigenic relatedness and leads to cross-reactive antibodies that predominantly target epitopes in S2. Conversely, antibodies targeting S1 and RBD are generally more type-specific^7–9^ and tend to have high neutralization potential. The differential homology across S1 and S2 domains affects the balance between neutralizing and other antibodies upon heterologous viral exposures^10^. Consequently, pre-existing sHCoVs immune responses impacted the development of SARS-CoV-2 immunity upon SARS-CoV-2 infection or COVID-19 vaccination. Notably, upon primary SARS-CoV-2 infection at the beginning of the pandemic, when the general population was immunologically naïve, the magnitude of boosted OC43 S2-specific IgG responses in certain individuals positively correlated with COVID-19 severity and these cross-reactive antibodies neutralized neither SARS-CoV-2 nor OC43^11,12^. This observation raised concerns on immunopathological interactions between SARS-CoV-2 and OC43 and supported a role for original antigenic sin (OAS) in severe COVID-19^10,13^. OAS refers to a negative attribute of immune memory whereby the immune system preferentially recalls antibodies against a previously encountered virus which is antigenically related, rather than generating a more effective virus-specific response. In contrast, recent infection with OC43 has been correlated with reduced SARS-CoV-2 seroconversion rates, suggestive of cross-protection, although this was not related to OC43 S-specific IgG responses^14^. The role of OC43 in enhancing or compromising immune protection against COVID-19 remains unclear in the increasingly immune population over the course of the pandemic.

In addition, little is known about the impact of the immune interactions between sHCoVs and SARS-CoV-2 in COVID-19 convalescent individuals, as well as their impact on protection against sHCoVs, and if this changed over the course of the pandemic^15–18^. A deeper understanding of cross-reactive immune responses between sHCoVs and SARS-CoV-2 can provide insights into the antigenic relations between coronaviruses as well as host responses. This is crucial for informed development of targeted vaccines and antibody-based immunotherapies for coronaviruses, including SARS-CoV-2.

Here, we investigated the magnitude, cross-reactivity over different domains and function of sHCoVs and SARS-CoV-2 directed antibodies in samples obtained from blood donors and healthcare workers (HCW) before and at the end of the pandemic. These were sampled between 2018 and 2019 (pre-pandemic) or in February and March 2023 (end-pandemic). We explored differences in the serum IgG and IgA titers, clonal antibody cross-reactivity patterns between sHCoV and relevant dominant SARS-CoV-2 variants, and their specificity towards the S1 and S2 domains. Additionally, we determined virus neutralization titers of serum and clonal antibodies against OC43 as a measure of antibody functionality and immune protection.

## METHODS

### Study design and clinical specimens

Blood samples were either collected as surplus material from blood donations at the Sanquin Blood Bank, Rotterdam, the Netherlands, as part of the SWITCH-ON study, a multicenter, open-label clinical COVID-19 vaccine trial (ClinicalTrials.gov ID: NCT05471440) and the COVA biobank study (**Supplementary Table 1**)^19^. The study protocols were approved by the Medical Ethics Committee of the Erasmus University Medical Center, Rotterdam, The Netherlands (SWITCH-ON: MEC-2022-0462; COVA: MEC-2014-398) and were conducted in accordance with the Declaration of Helsinki. Study participants were either adult voluntary blood donors (8 donors pre-pandemic and 8 end-pandemic) or HCW enrolled in the COVA biobank study (9 donors pre-pandemic) or the SWITCH-ON COVID-19 vaccination study (21 donors end-pandemic). Samples from the pre-pandemic group were collected between 2018 and 2019, prior to the beginning of the COVID-19 pandemic (n=17 plasma and n=8 plasma with paired PBMC samples). Samples from the end-pandemic group were collected in February or March 2023 (n=25 plasma; n=8 plasma with paired PBMC samples), at the end of the pandemic. Plasma and peripheral mononuclear cells (PBMC) were isolated from buffy coats, EDTA or heparin blood samples using Lymphoprep density-gradient (Stemcell Technologies) according to the manufacturer’s instructions. Plasma and PBMC samples were cryopreserved in aliquots at −20°C and −196°C for subsequent analysis, respectively.

### Protein microarray

For multiplex serology and B-cell analysis, a custom protein microarray (PMA) was generated as previously described^11^. The PMA consisted of a panel of S-trimer, S1, S2 and selected RBD antigens from the ancestral SARS-CoV-2 strain and relevant dominant variants (Delta, Omicron BA.1 and Omicron BA.5), as well as S-trimer and the S1 from the four endemic sHCoVs (229E, NL63, HKU1 and OC43), in addition to the S2 of HKU1 and OC43. 229E S1 reactivity was not determined for all donors because this antigen did not pass quality control. All antigens were obtained commercially (SinoBiological) except for the S-trimer antigens of sHCoVs, which were generated in house as previously described (**Supplementary Table 2**)^17^.

### Serology

Plasma IgG and IgA binding titers were determined against all antigens on the PMA as previously described^11^. In brief, PMA slides were blocked with Blotto (ThermoFisher Scientific) for 1 hour at 37°C and washed with PBS + 0.01% Tween (Sigma Aldrich; PBST). Four-fold dilution series of plasma were prepared in Blotto, starting at 1:10 and up to 1:163,840 and loaded onto the blocked and washed PMA slides for 1 hour at 37°C. Subsequently, slides were washed 4 times with PBST and stained with Alexa647-conjugated AffiniPure Goat anti-Human IgG Fc*γ* specific antibody (Jackson ImmunoResearch Laboratories; 109-605-008) or Cy3-conjugated AffiniPure Goat anti-Human Serum IgA *α*-chain specific antibody (Jackson ImmunoResearch Laboratories; 109-165-011) in Blotto for 1 hour at 37°C. After staining, slides were washed 4 times with PBST, once with ddH2O and airdried. PMA slides were scanned in a PowerScanner (Tecan). Serum IgG and IgA titers were determined as the EC50 using a 4-parameter logistic regression with GraphPad Prism v10.

### B-cell profiling

Short-term B-cells were cultured at limiting density to ensure clonal reactivity as described previously^11,20^. Briefly, B-cells were isolated from cryopreserved PBMCs using EasySep Human CD19 Positive Selection Kit (STEMCELL Technologies) according to manufacturer’s instructions. Cultures of 400 B-cells per well (pre-pandemic) or 50-100 B-cells per well (end-pandemic) were seeded in 96-well U-bottom plates in AIM-V AlbuMAX medium supplemented with 10% FCS, penicillin-streptomycin-glutamate (Invitrogen, ThermoFisher Scientific), and β-mercaptoethanol (Invitrogen) (B-cell medium). B-cell cultures were stimulated in a B-cell receptor-independent manner for 72 hours with 50 U/mL IL-2 (Novartis), 10 ng/mL IL-10 (Peprotech), 25 ng/mL IL-21 (Peprotech), 1 μg/mL resiquimod (Invivogen), and 10 times more 40 Gy x-ray irradiated, growth arrested L-CD40L cells (provided by J. Banchereau, The Jackson Laboratory for Genomic Medicine, Farmington, Connecticut, USA) than the number of CD19^+^ B-cells. CD40L expression and the absence of mycoplasma were confirmed for L-CD40L cells. After 72 hours, B-cells were cultured in B-cell medium supplemented with 25 ng/mL IL-21 for up to 14 days. Finally, culture supernatants were stored at −20°C until the measurement of IgG and IgA reactivity using PMA.

Culture supernatants were mixed 1:1 with Blotto and reactivity was analyzed using PMA. Bound IgG and IgA were stained fluorescently, analogous to serology. The mean fluorescent intensity (MFI; range 0 – 65350) of Alexa647-conjugated AffiniPure Goat anti-Human IgG Fc*γ* specific antibody (Jackson ImmunoResearch Laboratories; 109-605-008) and Cy3-conjugated AffiniPure Goat anti-Human Serum IgA *α*-chain specific (Jackson ImmunoResearch Laboratories; 109-165-011) was measured using a Powerscanner. A cut-off of 1000 MFI was set for all antigens, except for NL63 S1. This antigen gave higher background which was corrected by setting a batch-dependent cut-off (3000 for batch 1 and 6000 for batch 2, respectively) based on the MFI of non-reactive cultures (**Supplementary Figure 1**).

The frequency of reactive B-cells was calculated by counting the number of reactive wells divided by the total number of wells screened, multiplied by the number of cells per well. The frequency of reactive wells was used to calculate the probability of monoclonality (clonal accuracy) based on the Poisson distribution^21^.

### OC43 propagation and neutralization assay

The OC43 strain VR-1558 (ATCC, CCL-244) was propagated in MRC-5 cells (ATCC, CCL-171). Briefly, cells were infected at a multiplicity of infection (MOI) of 0.01. Progeny viruses were harvested six days post-infection and concentrated at 20,000 × *g* for 1.5 hour at 4°C through a 10% sucrose cushion.

Focus Reduction Neutralization Test (FRNT) assays were performed in A549 cells (ATCC, CCL-185). Plasma samples were heat-inactivated for 30 minutes at 56°C, then 3-fold serially diluted in 60 μL of Opti-MEM I (1×) + 1% penicillin-streptomycin (Gibco), starting at 1:20 in a 96-well V-bottom plate. Alternatively, B-cell culture supernatants were mixed 1:1 with Opti-MEM. Approximately 500 or 100 OC43 infectious viral particles in a volume of 60 μL were subsequently added to the diluted plasma or B-cell culture supernatant samples and incubated for 1 hour at 37°C, respectively. The virus-plasma or virus-supernatant mixtures (100 μL) were then transferred to confluent monolayers of A549 cells and incubated overnight at 37°C. Cells were fixed in 4% formalin, permeabilized with 70% ethanol, and blocked in 0.6% bovine serum albumin (BSA, Sigma) in PBS for 30 minutes. Cells were then incubated with rabbit anti-OC43 nucleocapsid antibody (Sino biological, 40643-762, 1: 2000 in 0.1% BSA), followed by incubation with Alexa Fluor 488-conjugated goat anti-rabbit IgG (Invitrogen, A32731, 1: 2000 in 0.1% BSA). Imaging was performed using the Amersham Typhoon Biomolecular Imager. Infected cells were quantified with ImageQuantTL 8.2.0.0 (GE Healthcare). The 50% foci reduction plasma titers (FRNT50) were calculated using a 4-parameter logistic regression with Graphpad Prism v10. For B-cell culture supernatants, the number of foci were compared directly. The lowest number of foci observed among the OC43 non-reactive supernatants was used as a cut-off distinguish neutralizing from a non-neutralizing supernatant. The respective culture media and pooled intravenous immunoglobulin preparations from healthy blood donors were included in all assays as negative and positive controls, respectively (data not shown).

### Statistics

Mann-Whitney tests were used to compare serology titers and reactive B-cell frequencies targeting sHCoV antigens and to compare OC43 neutralization titers between the pre- and end-pandemic groups. Differences in SARS-CoV-2 titers and frequencies were not statistically analyzed. For supernatants, differences in OC43 neutralization compared to the negative control group were analyzed using ANOVA. For correlation analyses, Pearson coefficient was calculated. All statistical analyses were carried out using GraphPad Prism v10 (GraphPad Prism Software).

## RESULTS

### Clinical characteristics of the cohorts

Pre-pandemic samples were retrieved between 2018 and 2019 (n=17 plasma and n=8 plasma with paired PBMC samples), while end-pandemic samples were obtained between February and March (n=25 plasma; n=8 plasma with paired PBMC samples). No significant differences were found in sex, nor in age of the individuals with available information from the different groups (**Supplementary Table 1**).

### Serum antibody titers against NL63 S1 and OC43 and HKU1 S2 were higher in end-pandemic donors

Serum IgG and IgA binding titers against seasonal sHCoVs and SARS-CoV-2 in pre- and end-pandemic donors were compared. Most pre-pandemic donors showed IgG titers against all four sHCoVs S-trimer and S1, as well as S2 domains of HKU1 and OC43. Interestingly, three out of 17 pre-pandemic donors (18%) had low, but detectable IgG titers against SARS-CoV-2 S2 (**Figure 1A**). End-pandemic donors were all seropositive for the four sHCoVs, and SARS-CoV-2 S-specific IgG titers were dominant over sHCoV across all S domains. Titers against sHCoV S-trimer were significatively higher in end-pandemic donors for NL63, HKU1 and OC43. For NL63, this was matched with increased titers against the S1, and for HKU1 and OC43 with increased S2 titers (**Figure 1A**). A similar trend was observed for IgA, were SARS-CoV-2 titers dominated in end-pandemic donors, albeit at lower magnitude than the observed for IgG. Moreover, end-pandemic donors showed significantly higher HKU1- and OC43-S2 titers (**Figure 1B**).

**Figure 1.**
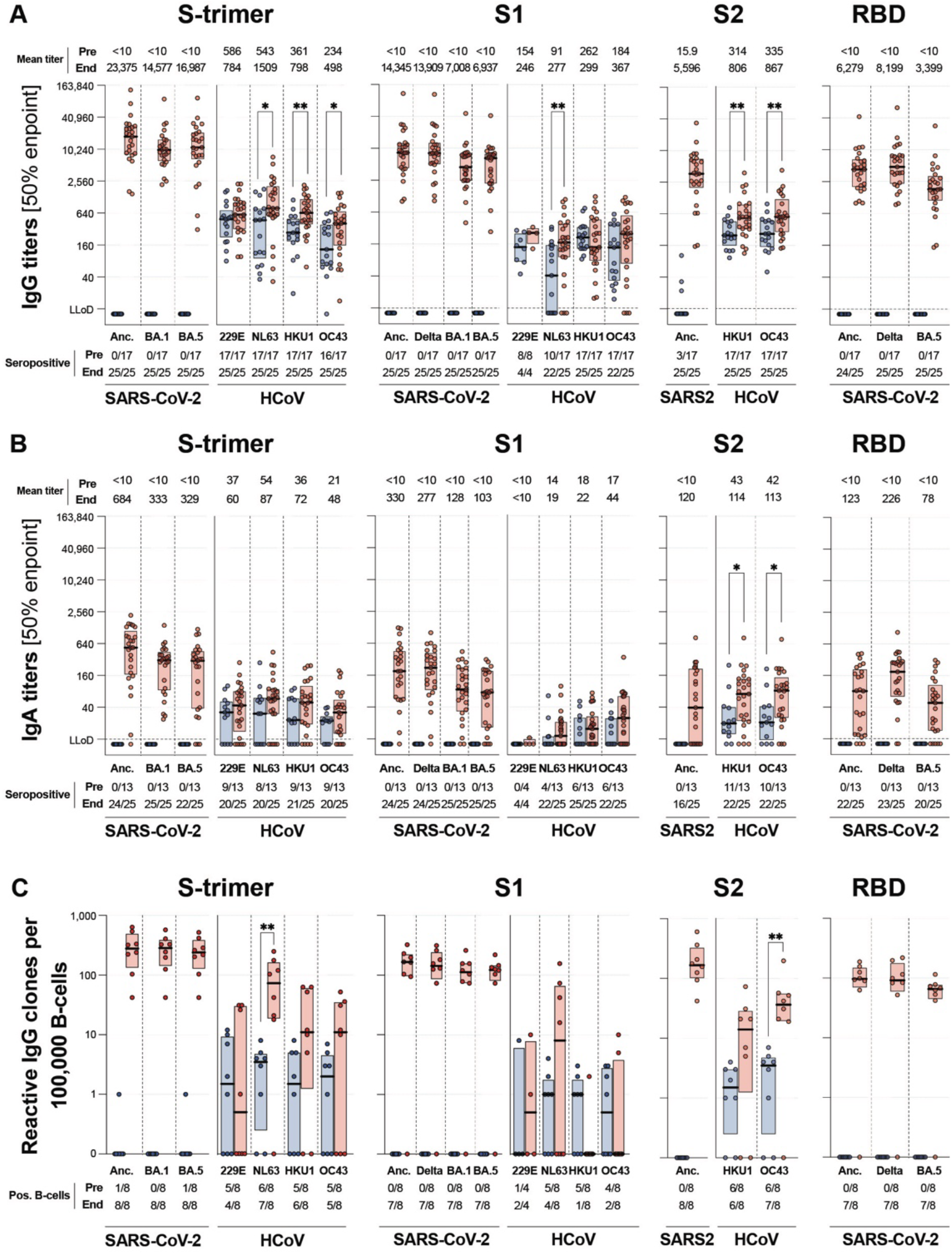
Serum IgG and IgA antibodies and frequency of IgG B-cells to seasonal human coronaviruses and SARS-CoV-2. **A**) Serum IgG and **B**) IgA 50% endpoint antibody titers towards prefusion stabilized spike (S-trimer), the S1 and S2 subdomains of spike and the receptor binding domain (RBD) of the ancestral SARS-CoV-2 strain (Anc.) and delta, omicron BA.1 and BA.5 variants and the seasonal human coronaviruses (sHCoV) 229E, NL63, HKU1 and OC43 were determined using protein microarray from healthy individuals sampled before (blue) or at the end of the pandemic (red). Mean titers are indicated on top and the number of seroconverted individuals is indicated below the graph. The lower limit of detection (LLoD) is indicated by a dotted line. **C**) The frequency of reactive IgG B-cells was determined by B-cell profiling for 8 donors sampled pre- (blue) and 8 donors sampled end-pandemic (red). Center line, median; boxes, upper and lower quartiles; significance of titer differences and B-cell frequencies for sHCoV were calculated using a Mann-Witney test and shown as \**p*<0.05 or \*\**p*<0.01.

### Frequencies of NL63 S-trimer and OC43 S2-reactive B-cells were higher in end-pandemic donors

The frequency of B-cell clones specific for the different coronavirus antigens was determined through B-cell profiling. The certainty of having a single reactive clone per well (clonal accuracy) per donor was high (average 91%, range 74-99%). Clonal accuracy was higher in pre-pandemic donors (97% ±2%; average ±SD) compared to end-pandemic donors (86% ±8%) due to the high frequency of S-trimer-specific clones in the latter group. Due to the lower frequency of detected S1, S2 and RBD-reactive clones, clonal accuracy for these antigens was above 95% for all donors.

In line with serological observations, in pre-pandemic donors we mostly detected sHCoV-reactive B-cells. However, rare SARS-CoV-2 ancestral and variant-specific IgG B-cells towards S-trimer were also detected in two donors. Although all donors were seropositive for sHCoV, circulating sHCoV S trimer, S1- and S2-reactive B-cells were not detected in part of them (**Figure 1C**). Nevertheless, the magnitude of the overall frequencies of antigen-specific circulating B-cells was in line with serum antibody titers (**Figure 1A/B**). In correspondence with increased serum titers, all end-pandemic donors consistently had high frequencies of SARS-CoV-2-reactive B-cells. Moreover, the frequencies of NL63 S-trimer and OC43 S2 specific IgG B-cells were higher compared to pre-pandemic donors. However, the frequencies in NL63 S1-specific IgG B-cells were not significantly different (**Figure 1A/C**).

### Cross-reactive IgG to SARS-CoV-2 and sHCoV were only detected in end-pandemic donors

To study the nature of the higher frequency of sHCoV-specific B-cells in end-pandemic donors, we analyzed the cross-reactivity patterns of reactive IgG and IgA clones, stratified for each set of S trimer, S1, S2 and RBD antigens (**Figure 2**). In pre-pandemic donors, 74 IgG (**Figure 2A**) and 29 IgA (**Figure 2B**) reactive clones to the S trimer of the different coronaviruses were detected, of which only 3 IgG and 2 IgA clones were reactive to SARS-CoV-2.

**Figure 2.**
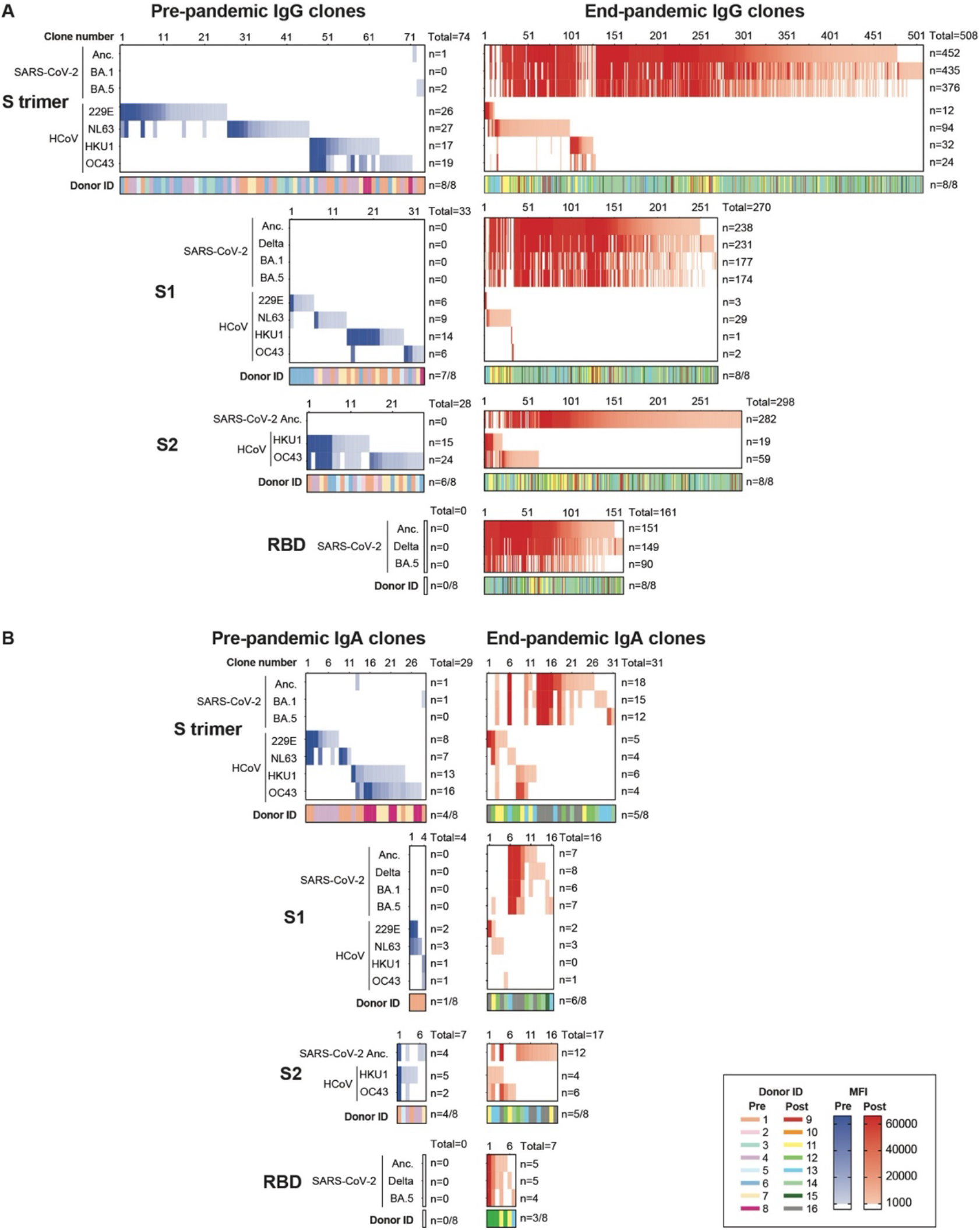
Cross-reactivity of individual IgG and IgA B-cell clones towards SARS-CoV-2 and seasonal human coronaviruses. **A**) The clonal IgG cross-reactivity patterns of CD19^+^ B-cells cultured at limiting concentration was determined using B-cell profiling. The mean fluorescent intensity of each clone for each antigen is indicated in pre-pandemic (blue shades) and end-pandemic (red shades). The clone number, total number of reactive clones per antigen, as well as the color-coded donor ID and number of donors with reactive clones are indicated. **B**) Clonal cross-reactivity patterns of IgA clones are plotted similarly.

In end-pandemic donors, 508 IgG and 31 IgA S-trimer reactive clones were detected, of which the majority were reactive and specific to SARS-CoV-2. Strong cross-reactivity between ancestral SARS-CoV-2, delta and the omicron BA.1 and BA.5 variants was detected across S trimer, S1 and RBD. The level of cross-reactivity between alpha and beta-sHCoVs was limited, yet some cross-reactive IgG and IgA clones were detected between these two genera (**Figure 2**). Strikingly, a substantial proportion of SARS-CoV-2-reactive IgG and IgA clones were cross-reactive with sHCoV. NL63 S1-reactive IgG clones, as well as HKU1 S2- and OC43 S2-reactive IgG and IgA clones showed cross-reactivity towards the respective SARS-CoV-2 antigen (**Figure 2**).

### End-pandemic donors showed a shift in immunodominance towards OC43 S2

To further delineate the extent of these immune interactions, we quantified the number of IgG clones that react or cross-react with SARS-CoV-2 and alpha- and beta-sHCoV separately. For alpha-sHCoV, we split the analysis for S trimer and S1 and for beta-sHCoV, we split the analysis for S trimer, S1 and S2 (**Figure 3A**). The higher numbers of NL63 S-trimer and S1-reactive IgG clones, and OC43 S2-reactive clones detected end-pandemic were mostly explained by the high number of clones that cross-recognized SARS-CoV-2 and their sHCoV counterpart antigen. In total, 20 cross-reactive clones targeting the S1 of SARS-CoV-2 and NL63 and 46 clones targeting the S2 of SARS-CoV-2 and OC43 were detected end-pandemic (**Figure 3A**).

**Figure 3.**
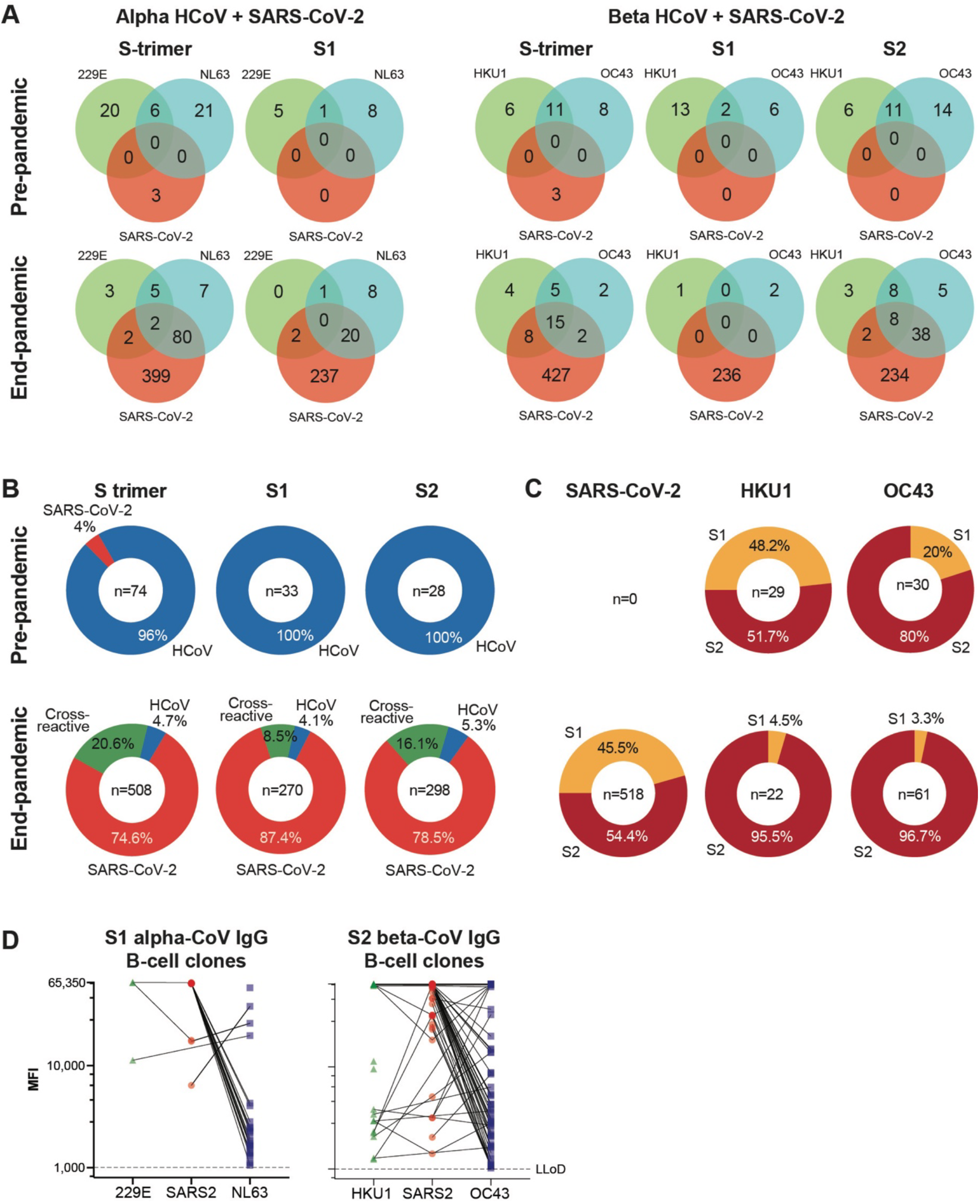
Quantification of reactive IgG B-cell clones. **A**) Type specific and cross-reactive clones targeting alpha-CoV and SARS-CoV-2 or beta-CoV and SARS-CoV-2 pre- and end-pandemic are enumerated and plotted in Venn diagrams for S-trimer, S1 and S2. **B**) The fraction of clones that are SARS-CoV-2-specific (red), sHCoV-specific (blue) or cross-reactive (green) from pooled pre- and pooled end-pandemic donors are plotted. **C)** Proportion of beta-CoV clones, including SARS-CoV-2, that are reactive towards S1 (orange) or S2 (dark red) pre- and end-pandemic. **D**) The level of reactivity of all 229E, NL63 and SARS-CoV-2 S1- (left) and HKU1, OC43 and SARS-CoV-2 S2- (right) reactive IgG clones detected in end-pandemic donors is indicated as the mean fluorescent intensity (MFI; range: 1000-65350). Cross-reactivity of clones is indicated by a connecting line.

This increased cross-reactivity was also reflected in their relative abundance. In pre-pandemic donors, 71/74 (96%) of all S trimer-specific IgG clones targeted seasonal sHCoV. In end-pandemic donors, SARS-CoV-2-reactive IgG clones accounted for 95.2% of the total coronavirus-specific response, 20.6% of which were cross-reactive with sHCoV and 4.7% selectively reacted with sHCoV (**Figure 3B**).

The high cross-reactivity impacted the relative immune dominance of S1 and S2 of beta-sHCoV. In pre-pandemic donors, the proportion of clones targeting S2 was 51.7% for HKU1 and 80% for OC43. In end-pandemic donors, SARS-CoV-2 reactive clones were evenly distributed between S1 (45.5%) and S2 (54.4%). However, because of the high proportion of cross-reactive clones targeting S2, the relative frequency of S2 targeting clones was higher for HKU1 (95.5%) and OC43 (96.7%) end-pandemic (**Figure 3C**). Similar trends were observed for IgA clones (**Supplementary Figure 2**). Altogether, this suggests a shift in target recognition towards the S2-domain of beta-sHCoV in end-pandemic donors.

### SARS-CoV-2/sHCoV cross-reactive clones preferentially reacted with SARS-CoV-2

To gain insight in the binding preference for SARS-CoV-2 or sHCoV of cross-reactive IgG clones, we compared the MFI for each clone towards each of the respective antigen. For both NL63 S1 and OC43 S2, most cross-reactive clones showed higher MFI towards the SARS-CoV-2 antigen compared to the sHCoV antigen (**Figure 3D**). This suggests that out of all cross-reactive clones, most have higher binding affinity for SARS-CoV-2.

### OC43 serum neutralization titers were higher in end-pandemic donors

To address a potential functional impact of the altered recognition of OC43 due to the increased immunodominance of S2 observed in end-pandemic donors, we assessed the serum neutralization titers using a focus reduction neutralization test (FRNT). The OC43 neutralization titers were significantly higher in the end-pandemic donors, suggesting the detected S2-specific cross-reactive antibodies had neutralizing potential (**Figure 4A**).

**Figure 4.**
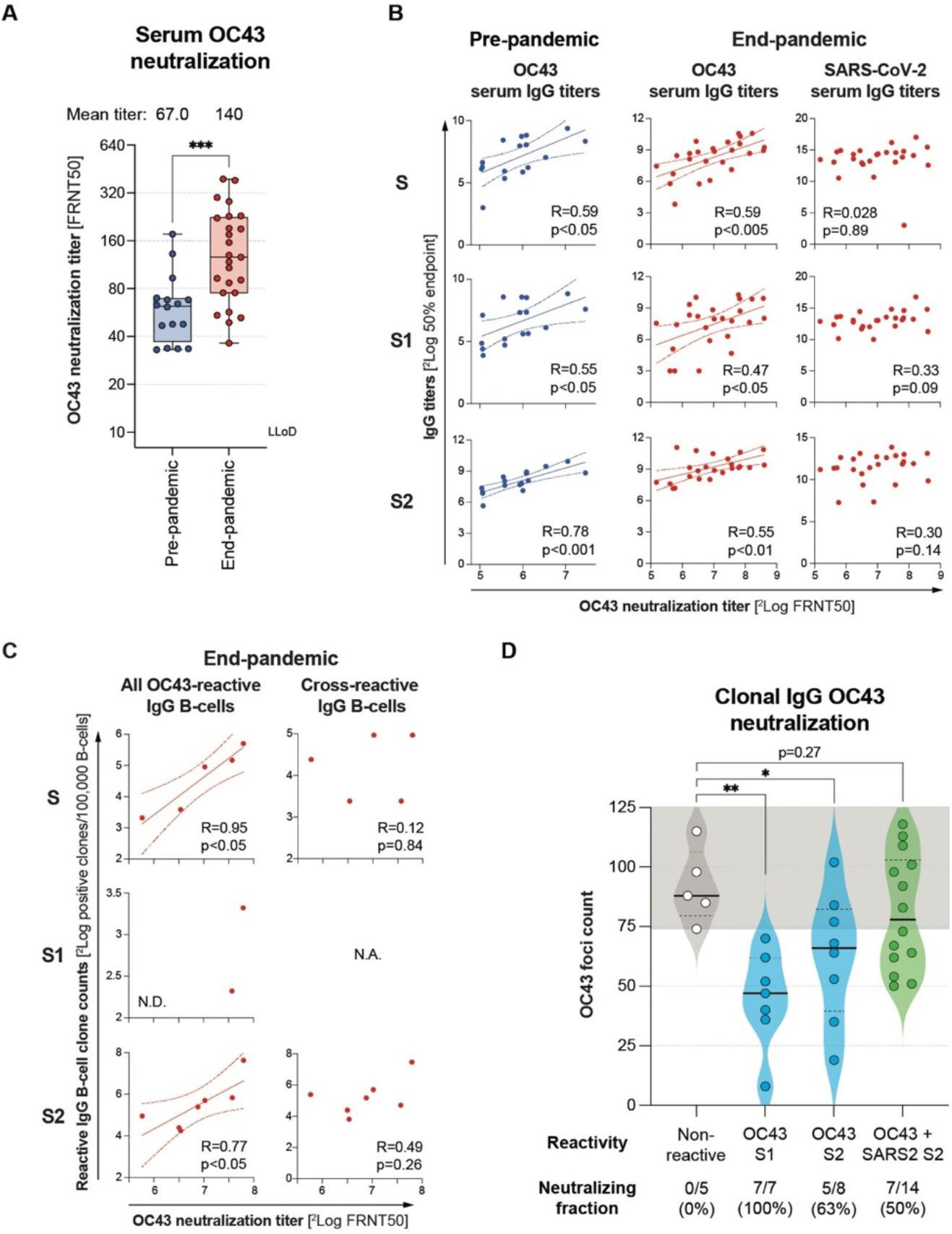
OC43 neutralization and correlations with serum IgG titers or B-cell clone counts. **A**) Serum OC43 neutralization potential was determined using 50% focus reduction neutralization tests (FRNT50) and compared between pre- (blue) and end-pandemic (red) donors. **B**) Linear regression analysis between serum OC43 neutralization titers and OC43 serum IgG binding titers (pre-pandemic, blue) or OC43 and SARS-CoV-2 serum IgG binding titers (end-pandemic, red). **C**) Linear regression analysis between serum OC43 neutralization titers and all OC43-reactive (left panels) or SARS-CoV-2/OC43 cross-reactive IgG B-cell clone counts (right panels) for S-trimer (top panels), S1 (middle panels) and S2 (bottom panels). All serum titers and reactive B-cell clone counts, normalized per 100,000 analyzed CD19+ B-cells were 2log transformed. **D**) IgG B-cell culture supernatants that were OC43 S1- or S2-specific, or SARS-CoV-2/OC43 S2-cross-reactive were tested in a OC43 focus reduction assay and compared with OC43 non-reactive controls. Significant differences were calculated by unpaired t-test and plotted as *p<0.05, **p<0.01 and ***p<0.001. The regression line and 95% confidence interval are plotted for significant correlations (*p*<0.05). N.A.: not applicable, N.D.: not determined.

### The frequency of OC43 S2-specific IgG B-cell clones correlated with OC43 neutralization in end-pandemic donors

To determine the relative contribution of OC43 and SARS-CoV-2 S-trimer, S1 and S2 IgG to OC43-neutralization, we correlated the serum IgG titers with OC43 FRNT50 titers. In both pre- and end-pandemic sera, OC43 neutralization titers correlated with OC43 S-trimer, S1 and S2 specific IgG titers. In contrast, correlations with SARS-CoV-2 serum IgG titers in end-pandemic donors were not significant (**Figure 4B**). Because only a minor fraction of the total SARS-CoV-2 clones cross-reacted with OC43 (46/235 clones; 19.6%, **Figure 2A and 3A/B**) such correlations may be difficult to identify at the polyclonal serum level.

Therefore, we subsequently assessed the contribution of OC43 S-trimer and S2 reactive IgG B-cell clones to serum OC43 neutralization by correlating the FRNT50 titers with IgG B-cell counts in end-pandemic donors. Due the limited number of pre-pandemic OC43-reactive and S1-reactive clones (**Figure 1C**) these correlations were only performed for S-trimer and S2 in end-pandemic donors. In end-pandemic donors, the frequency of S-trimer and S2-specific clones significantly correlated with serum OC43-neutralizing titers, supporting a contribution of OC43 S2-reactive IgG B-cells to the serum OC43 neutralizing response (**Figure 4C**). However, even though the majority of OC43 S2-reactive clones were SARS-CoV-2/OC43 S2-cross-reactive (46/59; 78%), in a sub-analysis with only the cross-reactive clones this correlation was not detected (**Figure 4C**). This could be related to lack of statistical power or to a minor contribution of the cross-reactive clones to the serum OC43 neutralization potential, compared to OC43 S2-specific IgG B-cell clones.

### OC43 S1 and S2-specific IgG clones showed OC43 neutralization potential

To investigate the OC43 neutralization potential of OC43 S1 and S2-specific, and SARS-CoV-2/OC43 S2-cross-reactive IgG clones further, we determined the OC43 neutralization potential of surplus culture supernatants using an adapted OC43 focus reduction assay. All OC43 S1 reactive clones (7/7; 100%) and 5 out of 8 (63%) OC43 S2-reactive clones showed a variable degree of OC43 foci reduction compared to non-reactive supernatants, which in grouped analysis was significant for S1 and S2(p<0.01 and p<0.05, respectively; **Figure 4D**). For SARS-CoV-2/OC43 S2-cross-reactive clones, 7/14 (50%) showed a reduction in the number of detected OC43 foci compared to the negative control, suggesting that part of the SARS-CoV-2/OC43 S2-cross-reactive clones has neutralization potential. However, in a group analysis this was not significantly different from non-OC43 reactive supernatants (p=0.27; **Figure 4D**), which may be caused by limited statistical power in this sub-analysis or heterogeneity in the neutralization potential of S2 cross-reactive clones. When analyzing OC43 S2-specific and SARS-CoV-2/OC43 S2-cross-reactive B-cells together, the reduction of OC43 foci was also not significant (p=0.22, data not shown), suggesting that overall, OC43 S2-specific clones have higher neutralization potential compared to SARS-CoV-2/OC43 S2-cross-reactive IgG. Nevertheless, together with the immunodominance of cross-reactive clones, these analyses support that part of the SARS-CoV-2/OC43 S2-cross-reactive IgG contribute to the higher serum OC43 neutralization observed in end-pandemic donors.

## DISCUSSION

The clinical impact of antigenic similarity between SARS-CoV-2 and sHCoVs on immune protection has been debated ^11,12,22–26^. There are indications for OC43-induced OAS in severe COVID-19 patients after primary SARS-CoV-2 infection early in the pandemic^11,12,23,25,26^. Yet, how these immune interactions have evolved over time in the general adult population, where the SARS-CoV-2 immune status has become increasingly diverse and where only a fraction suffered from severe COVID-19, is poorly understood. In this study, we retrospectively compared the immunorecognition of sHCoVs and SARS-CoV-2 and their interplay before and at the end of the COVID-19 pandemic in cross-sectionally sampled adult blood donors that either had no or presumably had multiple exposures to SARS-CoV-2 through vaccination and/or infection, respectively.

Compared to the pre-pandemic donors, we show higher NL63 S1, HKU1 S2 and OC43 S2 specific antibody titers in end-pandemic donors, which confirms previous findings^16–18,27^. In contrast to OC43, for NL63 and HKU1 no associations with severe COVID-19 have been described. Conversely, no aberrancies in medically attended or hospitalized sHCoV cases were reported^28^. These observations suggest that the net impact of the SARS-CoV-2 pandemic on the severity of infection with seasonal coronaviruses was neither significantly detrimental nor beneficial at the population level.

The antigenic relatedness between NL63 S1 and SARS-CoV-2 S1 is currently not clear. Distinct orientations of the C-terminal and N-terminal domains in S1 show overlapping structural features between both viruses^29^. This might in part be explained by the similar use of ACE-2 as an entry receptor^30^. The structural similarities in S1 could be targeted by cross-reactive antibodies with neutralization potential, although confirmation through structural and functional antibody analysis is required.

For HKU1, the level of sequence homology with SARS-CoV-2 within S2 is comparable with that of OC43^5,31,32^. Despite these similarities, with 10 out of 21 HKU1-reactive clones being cross-reactive with SARS-CoV-2 (48%), the number of detected SARS-CoV-2/HKU1 cross-reactive B cells was relatively low compared to OC43, where 46 out of 59 clones (78%) were cross-reactive. Previously, depletion of S2-directed antibodies in convalescent sera, yielded a significant reduction to the antibody binding of OC43 S, but had little effect on HKU1 S binding antibodies^17^. These discrepancies are likely related to limited affinity of cross-reactive antibodies. Taken together, this suggests a lower contribution of SARS-CoV-2 S2-directed antibodies in HKU1 S binding compared to OC43.

The detected higher OC43 S2 IgG titers and frequency of OC43 S2-reactive B-cells in end-pandemic donors is reminiscent of the observed boost of OC43 B-cells in severe COVID-19 patients^11,12^. However, while in severe COVID-19 patients the cross-reactivity of boosted OC43 S2-reactive towards SARS-CoV-2 was limited, we observed that SARS-CoV-2/OC43 S2-cross-reactive clones preferentially targeted SARS-CoV-2. This poses questions on the heritage and maturation of cross-reactive antibodies as OC43-induced clones may acquire reactivity towards SARS-CoV-2 by affinity maturation, or SARS-CoV-2 may induce clones that are by default cross-reactive or acquire this potential by affinity training after OC43 exposure^11^Although the preferential recognition of SARS-CoV-2 suggests that the S2-cross-reactive antibodies detected here were induced by SARS-CoV-2 infection or vaccination, claims on heritage and affinity maturation would need to be substantiated with longitudinal tracing of clones using BCR sequencing to address somatic hypermutations combined with antibody affinity measurements.

Functional differences between OC43-reactive antibodies after primary SARS-CoV-2 infection and/or COVID-19 vaccination are not yet well understood. COVID-19 convalescent and vaccinated individuals are both known to have increased OC43 binding titers^18,33,34^. When comparing convalescent and vaccinated donors, the first had higher OC43 antibody-dependent cellular cytotoxicity, complement deposition and phagocytosis^35^ compared to vaccinated-only donors^35^. In addition, vaccinated donors with a subsequent breakthrough SARS-CoV-2 infection had higher OC43 neutralizing activity in plasma and saliva upon recovery^16^, which is in line with what we observe in this study.

OC43 S1-specific IgG showed highest OC43 neutralization potential. However, we show that OC43 S1-specific serum IgG titers, B-cell frequencies including their cross-reactivity with SARS-CoV-2 were not significantly different between pre- and end-pandemic donors. Therefore, the higher serum OC43 neutralization titers are most likely explained by the higher OC43 S2-reactive IgG response. In turn, this aligned with the high level of SARS-CoV-2/OC43 S2-cross-reactivity in end-pandemic donors.

Differences in antibody functionality between groups may result from conformational differences in SARS-CoV-2 S. Whereas mRNA vaccination selectively elicits a response towards a proline-stabilized prefusion spike, infection may induce a response to a much wider range of S conformations that additionally expose epitopes within the fusion machinery located in S2. While certain epitopes accessible in an open or post-fusion conformation of S in S2 are targeted by non-neutralizing antibodies^36^, antibody binding epitopes in the pre-fusion stem-helix of SARS-CoV-2 S2 correlate with broad neutralization against beta-CoVs^3,37–40^ and are prevalent in convalescent vaccinated individuals but were scarce in vaccinated-only and convalescent-only individuals^9^. This highlights the functional heterogeneity of S2-directed antibodies depending on conformational target epitopes, which corresponds with the variable OC43 neutralization potential of S2-reactive clones that we observed.

Whether this leads to improved immune protection against OC43 infections remains to be determined. On the one hand, the dominance of SARS-CoV-2/OC43 S2 cross-reactive clones over OC43 S2-specific clones, combined with preferential binding of cross-reactive clones towards SARS-CoV-2 S2 observed in end-pandemic donors could lead to prozone effects, where an access of lower affinity cross-reactive antibodies outcompetes more potent type-specific responses. Indeed, our clonal OC43 focus reduction analysis suggested that the S2 cross-reactive IgG had lower binding and neutralization potential compared to OC43 type-specific IgG. Yet, the large number of cross-reactive clones may compensate for their lower potency. On the other, increasingly potent cross-reactive clones may be selected by repeated heterologous exposures and affinity maturation that have an increasingly beneficial role by providing broad immune protection. Therefore, continued longitudinal studies on the balance between type-specific and cross-reactive responses and how cross-reactivity evolves in the context of continued exposures to sHCoV and SARS-CoV-2 remain of interest.

Limitations of this retrospective, cross-sectional study design are the limited study size and the lack of paired pre-/end-pandemic samples, which restricts extrapolation of our results to the general population. Also, there was no clinical information on recent SARS-CoV-2 and/or sHCoV exposure of the study groups. The higher OC43 neutralization titers in end-pandemic donors could be a direct result of the induction of a SARS-CoV-2/OC43 cross-reactive response or by recent exposure to OC43. To investigate potential biases in OC43 exposure at a population level, we compared local epidemiological surveillance data^28^. Seasonal patterns of sHCoV detections were disturbed from the end of 2020 to early 2022 due to lock-down measures, yet circulation persisted at low levels. After that, both the number of detections and seasonal pattern reverted to similar levels observed pre-pandemic. Nevertheless, according to epidemiological data, end-pandemic donors were sampled between 10 and 15 weeks after the peak of reported sHCoV infections in the Netherlands, whereas pre-pandemic donors were more scattered. These observations indicate there is a potential risk of bias based on recent OC43 exposure (**Supplementary Figure 3**)^28^. In contrast, SARS-CoV-2 S1- and RBD-reactive antibodies decay faster than S2-directed antibodies upon infection for which an increased ratio of S1/S2-reactive antibodies is indicative of recent exposure^41,42^. Assuming similar differential kinetics for OC43 S1 and S2, the similarities in OC43 S1-directed antibody titers and frequencies compared to pre-pandemic donors, and increased S2-directed IgG response argues against recent OC43 exposure. Regardless of potential bias, recent OC43 infection would not explain the increased dominance of S2 over S1, and the increased cross-reactivity with SARS-CoV-2.

In conclusion, the unparalleled number of antibody clones analyzed here for their cross-reactivity potential and function offers a detailed and representative insight in coronavirus immune interactions. Our data shows that SARS-CoV-2 has altered immune recognition of NL63, HKU1 and OC43, due to antibody cross-reactivity. The increased IgG cross-reactivity with OC43 S2 and the higher levels of OC43 neutralization in end-pandemic donors are of particular interest. Despite the OC43-related immunopathogenic effect in severe COVID-19 donors observed early in the pandemic, our data on individuals sampled at the end of the pandemic indicate that SARS-CoV-2 S2-reactive B-cells and antibodies positively contribute to OC43 neutralization. Whether this translates into immune protection remains to be confirmed, but in that scenario, S2 might constitute a relevant target for the design of pan-corona vaccines.

## DATA AVAILABILITY

Upon a reasonable request, the corresponding author can supply the data in this study.

## ACKNOWLEDGEMENTS

This work was funded by the Horizon Europe project VEO (project number 874735) and RECoVER (101003589), the Dutch Research Council (NWO) project One Health Pact (109986), the NWO TKI-LSH Health~Holland project Punch-mAb (EMCLSH22024) and the Netherlands organization for health research and development (ZonMw) project Create2Solve Phase 1 (01142052310005).

## AUTHOR CONTRIBUTIONS

CGL, MAB and GPvN conceptualized the study. CGL, MAB, JZ, BB and PvdD performed experiments. MJvG and BLH provided critical reagents. ECvG, HMvdK, CGvK and RDdV provided clinical specimens. CGL, MAB and GPvN analyzed the data and generated the figures. CGL, MAB and GPvN wrote the manuscript. MAB, JZ, RDdV, CGvK, MJvG and MPGK reviewed and edited the manuscript. All authors critically reviewed and approved the manuscript.

