## Supplementary tables and figures for "SARS-CoV-2 cross-reactive B-cells outnumber seasonal coronavirus spike-specific clones at the end of the COVID-19 pandemic"

**Supplementary table 1.** Clinical characteristics of the cohorts.

| Cohort Characteristics | | | | | |
|---|---|---|---|---|---|
| **Pre-pandemic** | | | | | |
| **Study** | **Sex** | | **Age (years) ±SD** | **Serum n = 16** | **PBMC n = 8** |
| | **Female** | **Male** | | | |
| COVA Biobank n = 8 | 2 | 6 | 45 ± 5 | 8 | 0 |
| Blood bank n = 8 | N.D. | N.D. | N.D. | 8 | 8 |
| **End-pandemic** | | | | | |
| **Study** | **Sex** | | **Age (years)** | **Serum n = 25** | **PBMC n = 8** |
| | **Female** | **Male** | | | |
| SWITCH-ON n = 21 | 10 | 10 | 44 ± 6 | 21 | 4 |
| Blood bank n = 4 | N.D. | N.D. | N.D. | 4 | 4 |

Age and sex data from n=16 individuals obtained between 2018 and 2019 (pre-pandemic) and n=25 individuals obtained between February and March 2023 (end-pandemic) are shown. No difference was observed in age or sex composition of the groups. Data of blood bank donors were unavailable due to data protection policies; therefore, these were not determined (N.D.) and were excluded from the statistical analysis. SD; standard deviation.

**Supplementary Table 2.** Antigens on protein microarray.

| Virus (variant) | Antigen | Cat.No/UniProt ID | Supplier |
|---|---|---|---|
| SARS-CoV-2 (Ancestral) | S-trimer | 40589-V08H8 | SinoBiological |
| | S1 | 40591-V08H3 | SinoBiological |
| | S2 | 40590-V08B | SinoBiological |
| | RBD | 40592-V08H | SinoBiological |
| SARS-CoV-2 (Delta) | S1 | 40591-V08H23 | SinoBiological |
| | RBD | 40592-V08H90 | SinoBiological |
| SARS-CoV-2 (Omicrons BA.1) | S-trimer | 40589-V08H26 | SinoBiological |
| | S1 | 40591-V08H41 | SinoBiological |
| | RBD | 40592-V08H121 | SinoBiological |
| SARS-CoV-2 (Omicrons BA.5) | S-trimer | 40589-V08H32 | SinoBiological |
| | S1 | 40591-V08H46 | SinoBiological |
| | RBD | 40592-V08H130 | SinoBiological |
| HCoV-229E | S-trimer | NP_073551 | In-house[1] |
| | S1 | 40601-V08H | SinoBiological |
| HCoV-NL63 | S-trimer | AKT07952 | In-house[1] |
| | S1 | 40600-V08H | SinoBiological |
| HCoV-HKU1 | S-trimer | Q0ZME7 | In-house[1] |
| | S1 | 40021-V08H | SinoBiological |
| | S2 | 40021-V08B | SinoBiological |
| HCoV-OC43 | S-trimer | Q696P8 | In-house[1] |
| | S1 | 40607-V08H1 | SinoBiological |
| | S2 | 40607-V08B1 | SinoBiological |

All antigens were produced in HEK293 cells with a C-terminal His-tag, except S2 antigens. These were produced in Baculovirus-insect cells. [1]In-house proteins were produced as described in (Grobben et al., 2021a)

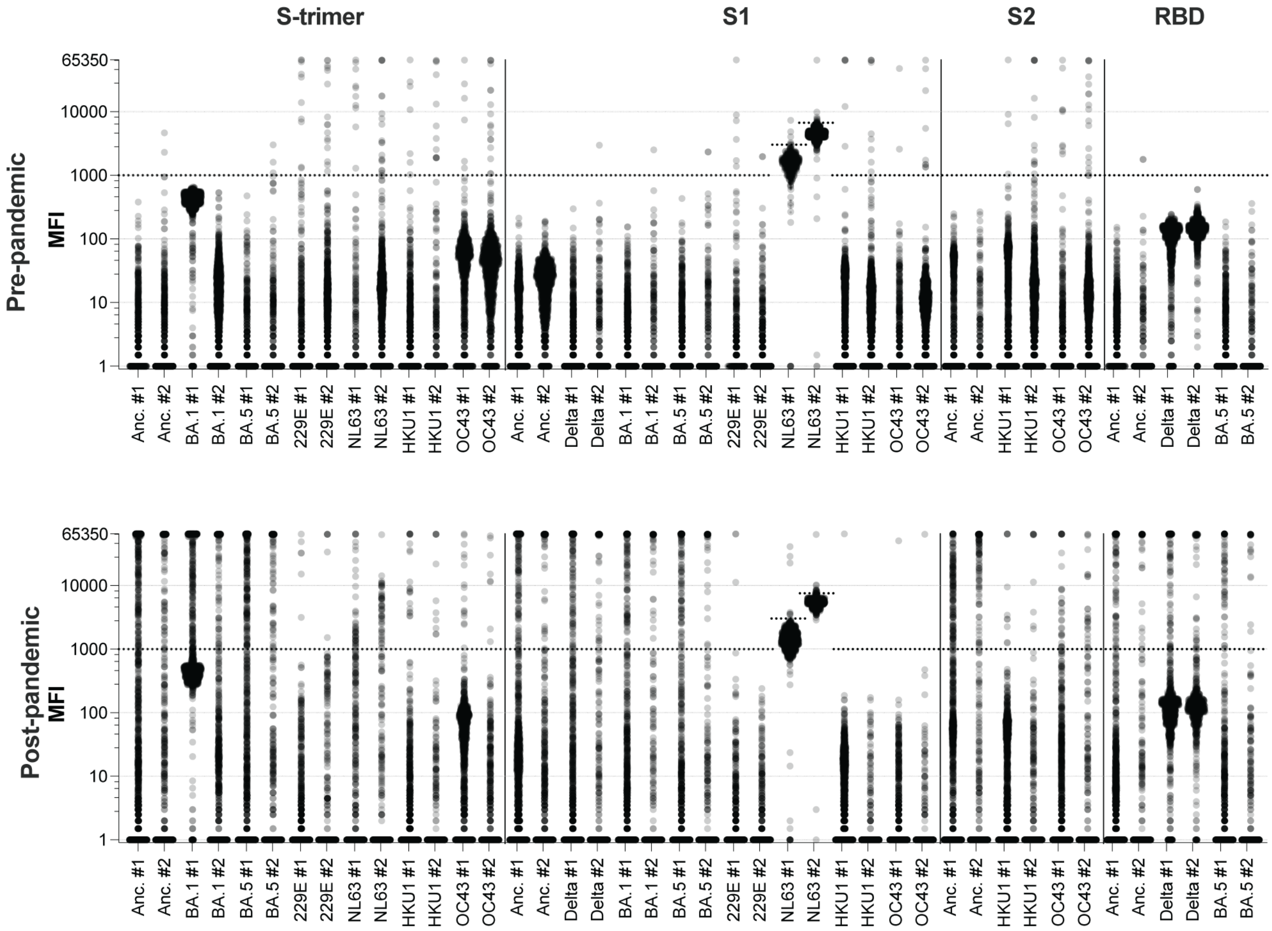


**Supplementary Figure 1. Background corrections for protein microarray per antigen.** The mean fluorescent intensity (MFI) of two antigen spots per antigen were plotted to define fluorescence cut-offs for positive cultures per antigen and per batch of slides (#1 and #2). A cut-off of 1000 MFI was set for all antigens, except for NL63 S1, due to a higher background signal. A batch-dependent cut-off was set for this antigen at 3000 MFI for batch #1 and 6000 MFI for batch #2 based on the MFI of non-reactive supernatants.

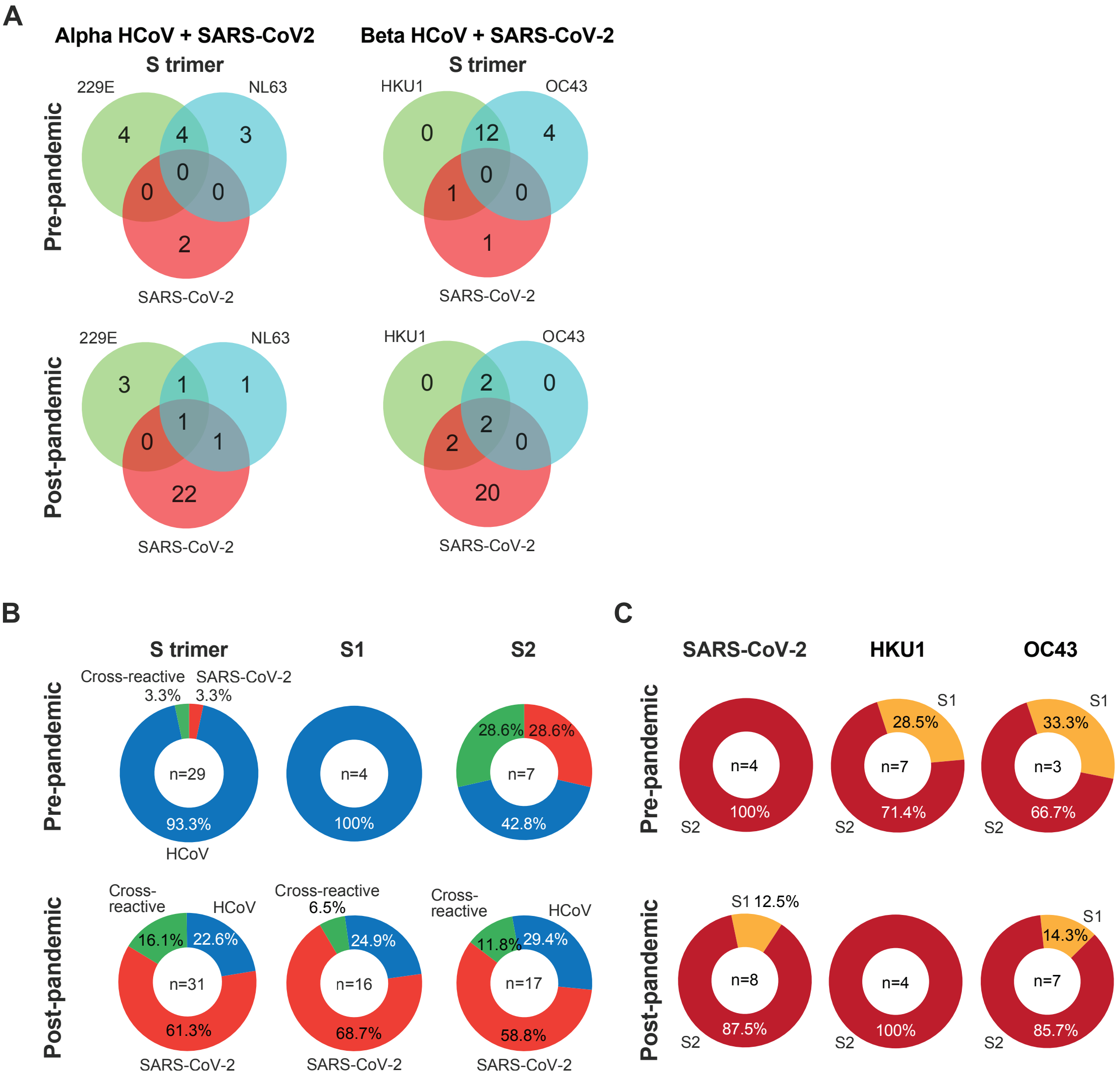


**Supplementary Figure 2. Quantification of reactive IgA B-cell clones. A**) Type-specific and cross-reactive clones targeting alpha-CoV and SARS-CoV-2 or beta-CoV and SARS-CoV-2 pre- and end-pandemic are enumerated and plotted in Venn diagrams. **B**) The fraction of clones that are SARS-CoV-2-specific (red), sHCoV-specific (blue) or cross-reactive (green) from pooled pre- (top panels) and pooled end-pandemic (bottom panels) donors are plotted. **C**) For beta-CoV and SARS-CoV-2 the proportion of clones that are reactive towards S1 (orange) or S2 (dark red) are indicated.

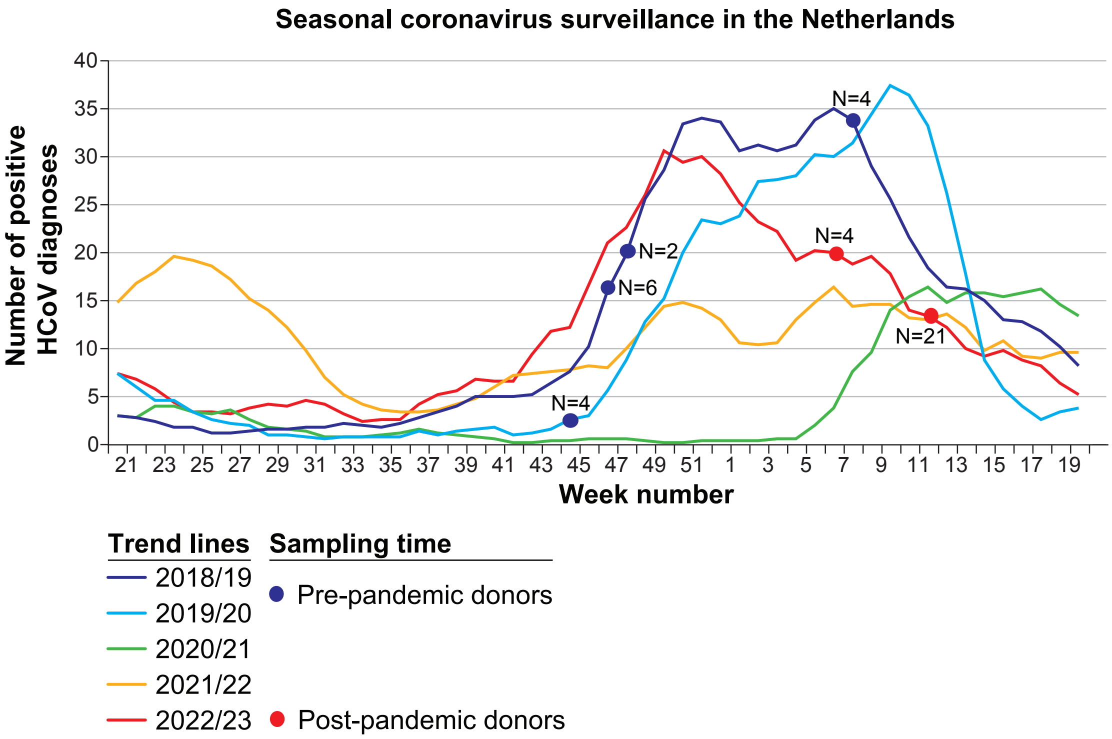


**Supplementary Figure 3. Seasonal coronavirus surveillance in the Netherlands.** 5-week moving average trendlines for the number of reported positive test results for seasonal coronaviruses (excluding SARS-CoV-2) in the virological laboratory surveillance in the Netherlands for the seasons 2018/19 up to 2022/23. Sampling time and number of donors included in our pre- and end-pandemic cohorts are indicated on the respective trendline. [Figure adapted from: *DFM Reukers et al. (2023), Annual report surveillance of acute respiratory infections in the Netherlands; winter 2022/2023*]
